# Identification of Shared and Unique Key Biomarkers of Alcohol Liver Cirrhosis and Non-Alcoholic Steatohepatitis Through Machine Learning Network-Based Algorithms

**DOI:** 10.1101/2024.10.17.24315623

**Authors:** Morteza Hajihosseini, Fernanda Talarico, Caroline Zhao, Scott Campbell, Daniel Udenze, Nastaran Hajizadeh Bastani, Marawan Ahmed, Erfan Ghasemi, Lusine Tonoyan, Micheal Guirguis, Patrick Mayo, Corinne Campanella

**Affiliations:** Applied Pharmaceutical Innovation, Edmonton, Alberta, T5J 4P6; School of Public Health, University of Alberta, Edmonton, Alberta, T6G 1C9; Faculty of Pharmacy and Pharmaceutical Sciences, University of Alberta, Edmonton, Alberta, T6G 2C8

**Keywords:** Liver, Cirrhosis, Steatohepatitis, Master Regulator, Key Driver

## Abstract

**Introduction:** Liver fibrosis can progress to cirrhosis, liver failure, or hepatocellular carcinoma, which often requires transplantation and burdens healthcare systems around the world. Advances in single-cell RNA sequencing and machine learning have enhanced the understanding of immune responses in many liver diseases particularly alcohol liver cirrhosis (ALC) and non-alcoholic steatohepatitis (NASH). This study aims to identify key biomarkers involved in these conditions and assess their potential as non-invasive diagnostic tools.

**Methods:** Two gene expression profiles GSE136103 and GSE115469 were used to conduct differential gene expression (DEG) analysis. Using the results from DEG analysis, we then applied two machine learning network-based algorithms, master regulator analysis (MRA) and weighted key driver analysis (wKDA), to identify potential biomarker genes for NASH and ALC.

**Results:** A total of 1,435 and 5,074 DEGs were identified for ALC and NASH compared to healthy controls, including 1,077 shared DEGs between the two diseases. The MRA showed HLA-DPA1, HLA-DRB1, IFI44L, ISG15, and CD74 as the potential master regulators of ALC and HLA-DPB1, HLA-DQB1, HLA-DRB5, PFN1, and TMSB4X as the potential master regulators of NASH. In addition, wKDA analysis indicated CD300A, FCGR2A, RGS1, HLA-DMB, and C1QA as the key drivers of ALC and INPP5D, NCKAP1L, RAC2, PTPRC, and TYROBP as key drivers of NASH.

**Conclusion:** This study presented a comprehensive framework for analyzing single-cell RNA-seq data, demonstrating the potential of combining advanced network-based machine-learning techniques with conventional DEG analysis to uncover actionable prognostic markers for ALC and NASH with potential use as target biomarkers in drug development.

## Introduction

Fibrosis is a common response to injury, marked by the excessive accumulation of extracellular matrix (ECM), which disrupts the restoration of connective tissues and impairs organ function. In the liver for example, this process raises the risk of cirrhosis. Inflammation and fibrosis are closely interlinked, as evidenced in many chronic liver diseases such as non-alcoholic fatty liver disease (NAFLD, a.k.a MASLD) (1,2), non-alcoholic steatohepatitis (NASH, a.k.a MASH) (3), and alcohol-associated liver diseases (ALD) (4). From 1990 to 2019, the overall pooled prevalence of NAFLD and NASH was 30.05% and 5.27%, respectively (5). North America and Australia regions had the highest prevalence of NAFLD and NASH in 2019 among all regions (5). In addition, the overall prevalence of ALD in the general population was 4.8% with variations from 1 to 16.1% reported in various studies (6).

Single-cell level RNA sequencing (scRNA-seq) analyses have been shown to be helpful in elucidating how injury signals, immune responses, and fibrotic pathways interact in liver diseases (7). Zhang et al. study on human single-cell mRNA-seq data focused on NASH tissues showed that immune response-related hub genes *CD53, LCP1, LAPTM5, NCKAP1L, C3AR1, PLEK, FCER1G, HLA-DRA* and *SRGN* were associated with NASH which was verified further in clinical samples and mouse models (8). Choudhary and Duseja’s review study revealed various marker genes associated with NAFLD and ALD (9). They showed that genes such as *ENPP1, IRS1, GCKR, PNPLA3, TM6SF2, MBOTAT, LIPIN1, AGTR1, SOD2*, and *KLF6* were associated with fibrosis in NAFLD patients, and *ADH, ALDH, PNPLA3, TM6SF2, MBOAT7, CTLA4*, and *IL1B* were connected with cirrhosis and hepatocellular carcinoma (HCC) in patients with ALD (9).

Advancements in sequencing technologies and bioinformatic tools have enhanced the diagnosis and treatment of various diseases (10–13), expanding opportunities to utilize large datasets and machine learning algorithms for personalized healthcare and improved patient care delivery. Several studies have employed machine learning algorithms to discover molecular signatures of liver diseases (14–17). Listopad et al. utilized logistic regression, k-nearest neighbours, and support vector machine methods to classify individuals with alcohol-associated hepatitis (AH), alcohol-associated cirrhosis, NAFLD, and healthy individuals using biospecimens collected from participants enrolled by the Southern California Alcoholic Hepatitis Consortium (14). Moreover, Shen et al. identified 12 key genes linked to the development and progression of HCC using mRNA expression profiles of three publicly available datasets and six machine learning algorithms, including AdaBoost, decision-tree, random tree, random forest, k-nearest neighbours, and Bayesian Net (17). However, studies that integrate machine learning strategies often rely on the gene expression profile of the disease and suffer from a lack of implementation of essential information from disease-associated networks. To address this issue, this study aims to find the key biomarkers for NASH and alcoholic liver cirrhosis (ALC) by applying a master regulator and weighted key driver algorithms on publicly available single-cell RNA datasets. Both machine learning and network-based algorithms consider disease-associated genes to identify candidate genetic biomarkers.

We performed differentially expressed gene (DEG) followed by the same analyses for different immune cell types. Subsequently, we used the results of DEG analysis for each disease to apply two machine learning algorithms and explore candidate biomarker genes for NASH and ALC. The results of this study will help in development of NASH and ALC treatments. Moreover, it shows the application of different bioinformatic tools, which can be applicable to various phases of the drug development process.

## Methods

The datasets utilized for analysis comprised 167,856 cells and 35,605 genes and were sourced from the Single Cell Portal (https://singlecell.broadinstitute.org/) for differential expression gene (DEG) and gene set enrichment analysis (GSEA). Metadata was accessible for 63,000 samples, with 8,928 of them being associated with liver diseases. Following the exclusion of cells with less than 1,000 reads (N=253) and those linked to primary biliary cholangitis (N=358), the study covered 8,317 cells. It specifically focused on cells attributed to alcoholic liver cirrhosis (ALC=1,953), non-alcoholic steatohepatitis (NASH=1,236), and healthy controls (5,128). The analytic dataset comprises samples from two studies, GSE136103 and GSE115469, which are available on Gene Expression Omnibus (GEO:http://www.ncbi.nlm.nih.gov/geo). Detailed information on the dataset and pre-processing steps has been previously described (18,19). Figure 1 shows the systematic process of data selection in this study.

**Figure 1.**
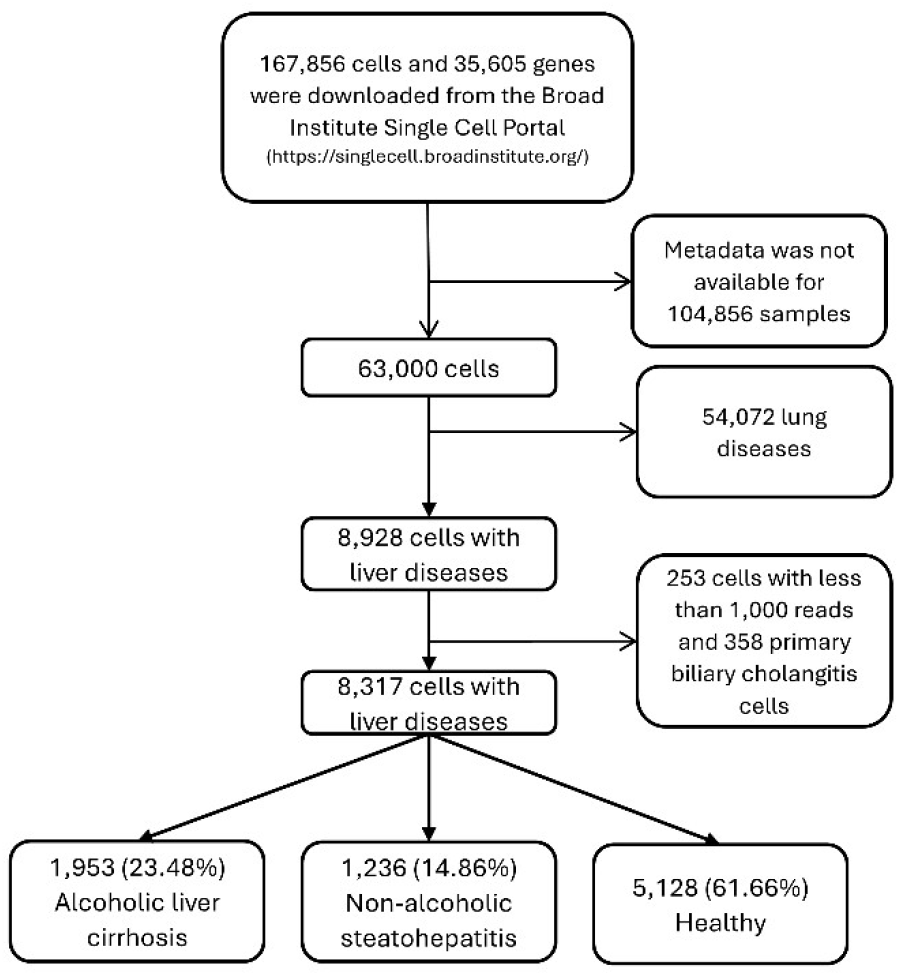
Flowchart describing the systematic process of data selection. This flowchart illustrates the steps taken to acquire, pre-process, and selectively filter data for inclusion in the study.

The metadata included ten different human cell types with their respective percentages: alveolar macrophages (4.7%), apoptosis-fated cells (0.3%), classical monocytes (23.4%), conventional dendritic cells (16.9%), Kupffer cells (6.9%), mature macrophages (28.5%), mature conventional dendritic cells (0.4%), mitotic cell cycles (0.3%), monocytes (8.7%), and non-classical monocytes (9.8%). We focused our cell-type-specific analyses on cell types with a frequency exceeding 5%.

### Differentially Expressed Gene (DEG) Analysis

We performed DEG analysis in *DESeq2 R Bioconductor* package v1.42.1 (20). In particular, we used the *DESeq* function to perform DEG analysis for ALC versus healthy and NASH versus healthy comparisons. The *DESeq* function calculates size factors and dispersion values and fits a negative binomial generalized linear model (GLM) to gene expressions. Additionally, we utilized the *lfcShrink* function with the *apeglm* option to estimate shrunken log2 fold changes and standard errors, using an adaptive *Student’s t* prior shrinkage estimator. This approach aims to preserve large differences and remove potential noise from the results (21). DEGs were selected using an adjusted p-value (q-value) threshold of <0.01. Venn diagrams were used to show the number of unique and shared DEGs for each comparison.

We conducted the same analyses for a specific group of cell types, which we referred to as the cell-type-specific DEG analysis. We then used the intersection of the top 20 ALC and NASH cell-type-specific DEG lists to identify the most important shared markers among and within cell types. Because of the smaller sample size in cell-type categories, we used a less conservative threshold of q-value<0.1 for the cell-type-specific DEG analysis. All heatmaps were sorted based on the average normalized gene expression from the lowest (blue) to the highest (red), and were created using the *ggplot2 R CRAN* package v3.5.0.(22).

### Exploring Candidate Biomarker Genes

We utilized two network-based pipelines to explore candidate biomarkers for each condition. *Corto* pipeline uses gene networks based on optimized pairwise correlations to find master regulator genes (23). The *Mergeomics* pipeline uses a network-based approach to find key drivers (hub genes) based on the disease associated genes (24,25).

### Master Regulator Analysis (MRA)

Master Regulator Analysis (MRA) was performed by comparing ALC versus healthy and NASH versus healthy tissues separately using the *corto* algorithm (23). We used the list of DEGs from each analysis in the previous step as centroids in the *corto* function to generate the regulon list. The regulon is a list of final centroids with at least one target. Then we normalized the gene expression data for ALC, NASH, and healthy tissues using variance stabilizing transformation (26) as recommended by the *corto* algorithm to obtain MRAs.

### Weighted Key Driver Analysis (wKDA)

The weighted key driver analysis (wKDA) was used to identify hub genes with network neighbourhoods that have an over-representation of disease-associated genes. The key driver concept is based on projecting disease-associated gene sets onto a network model of gene regulation that represents molecular interactions in the entire system. This analysis incorporates weights by considering the strength or reliability of the connections between genes, as represented by the network edge weights. We used the Mergeomics pipeline (24,25) and the embedded Bayesian Liver Network to find the top five key drivers based on the ALC and NASH DEG lists. In brief, the wKDA begins by searching a network for candidate hub genes with high connections. It then collects the neighbouring genes for each candidate hub gene and estimates the contribution of the disease-associated genes within the hub’s neighbourhood. If the contribution is stronger than expected by chance (i.e. significant p-value), it is concluded that the hub gene is a key driver of the disease-associated gene sets.

## Results

### Identification of DEGs in ALC and NASH tissues

In the entire sample, a total of 1,435 ALC-related DEGs and 5,074 NASH-related DEGs were identified, including 1,077 shared DEGs between the two conditions (for more details, see Figure S1 in the supplementary materials). Figure S2 shows the number of unique and shared DEGs for ALC and NASH in our cell-type specific DEG analysis.

The heatmaps in Figure 2 show the up-regulation and down-regulation of the top 20 DEGs. In Figure 2A, out of the top 20 DEGs in ALC, 6 genes (S100A11, NPC2, GPX1, HLADRB1, TYROBP, and MT-ND2) were highly upregulated in most cells, while HLA-DQA2 and ZNF90 were the most down-regulated. Figure 2B shows the results for NASH. There is a strong up-regulation of FTL followed by MT-CO1, MT-CO3, MT-CO2, MT-ND4, JUNB, MT-ATP6, MT-ND2, MT-ND3, S100A11, and HLA-DRB5, while CD5L, MARCO, ALB, CD163, and LIPA were down-regulated.

**Figure 2.**
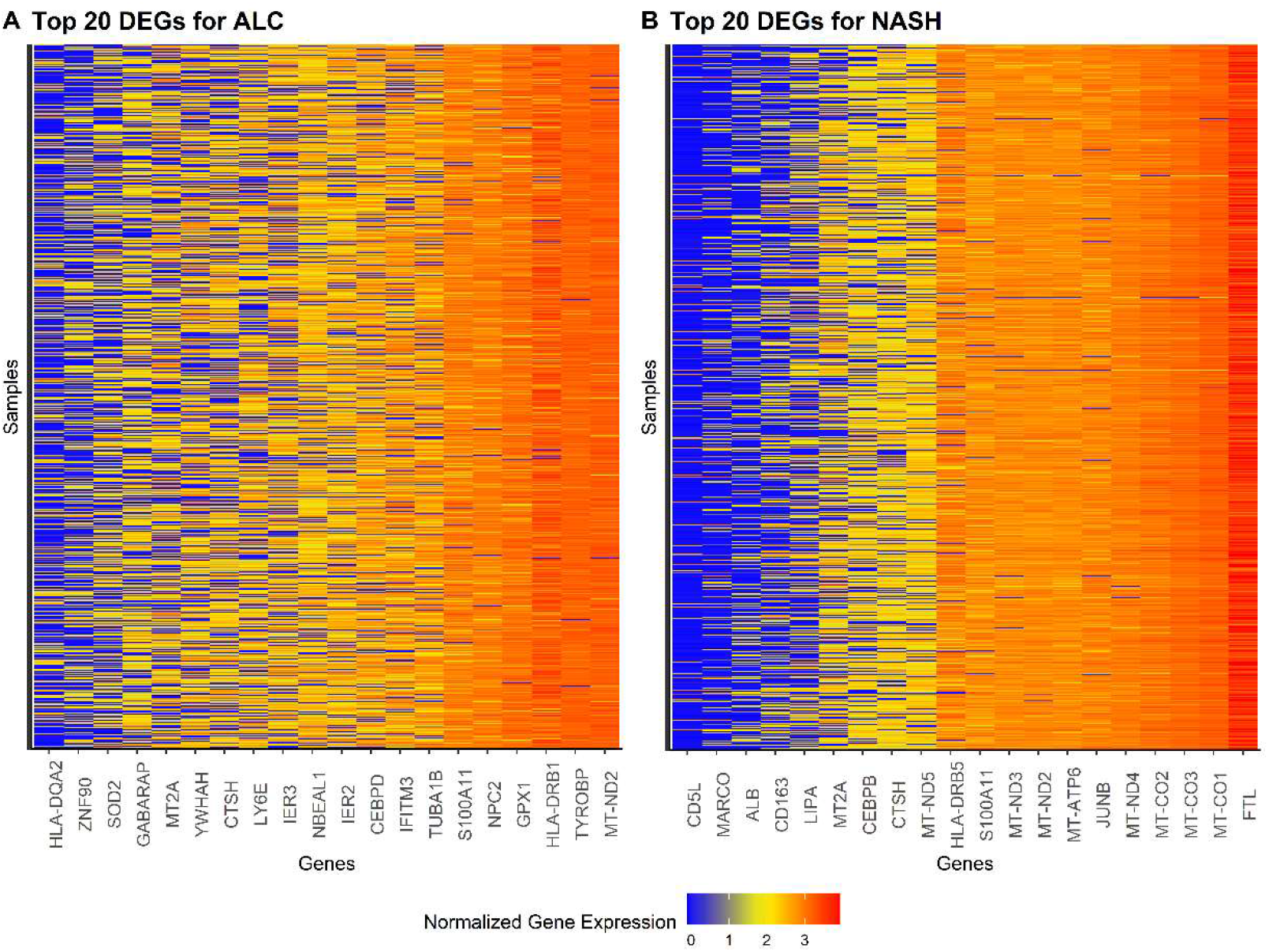
Heatmap of top 20 DEGs: (**A**) DEGs for ALC tissues. (**B**) DEGs for NASH tissues. Upregulated genes are marked in red and downregulated genes are marked in blue. All DEGs are sorted and normalized. DEGs=differentially expressed genes. ALC=alcoholic liver cirrhosis. NASH=non-alcoholic steatohepatitis.

The heatmaps in Figure 3 illustrate the up-regulation and down-regulation of cell-type-specific DEGs. In classical monocyte cells (Figure 3A), *MALAT1* is up-regulated, and *THBS1* is down-regulated in ALC, while *TMSB4X* is up-regulated, and *ARF6* is down-regulated in NASH (Figure 3B). Conventional dendritic cells (Figure 3C) show the up-regulation of *HLA-DRB1* and down-regulation of *S100A8* in ALC and the up-regulation of *HLA-DRB5* and down-regulation of *C1orf56* in NASH (Figure 3D). Additionally, Kupffer cells (Figure 3E) depict the up-regulation of CD74 and the down-regulation of HLA-DQA2 in ALC, while the up-regulation of *CCL4L2* and the down-regulation of *ARL4C* in NASH (Figure 3F). Macrophage cells (Figure 3G) exhibit the up-regulation of *TYROBP* in ALC and the up-regulation of *FTL*, while down-regulation of *CD5L* in NASH (Figure 3H). Furthermore, monocyte cells (Figure 3I) display the up-regulation of *FTH1* and the down-regulation of *HLA-DQA2* in ALC, while the up-regulation of *MT-CO1* and the down-regulation of *EIF5A* in NASH (Figure 3J). Non-classical monocyte cells (Figure 3K) illustrate the up-regulation of *FTH1* and the down-regulation of *HLA-DQA2* in ALC, while the up-regulation of *TMSB4X* and the down-regulation of *PABPC4* in NASH (Figure 3L). Additionally, Figure S2 in the supplementary materials shows the number of unique and shared cell-type-specific DEGs for ALC and NASH. Table S1 summarizes the list of the top 20 cell-type-specific unique and shared DEGs for each condition.

**Figure 3.**
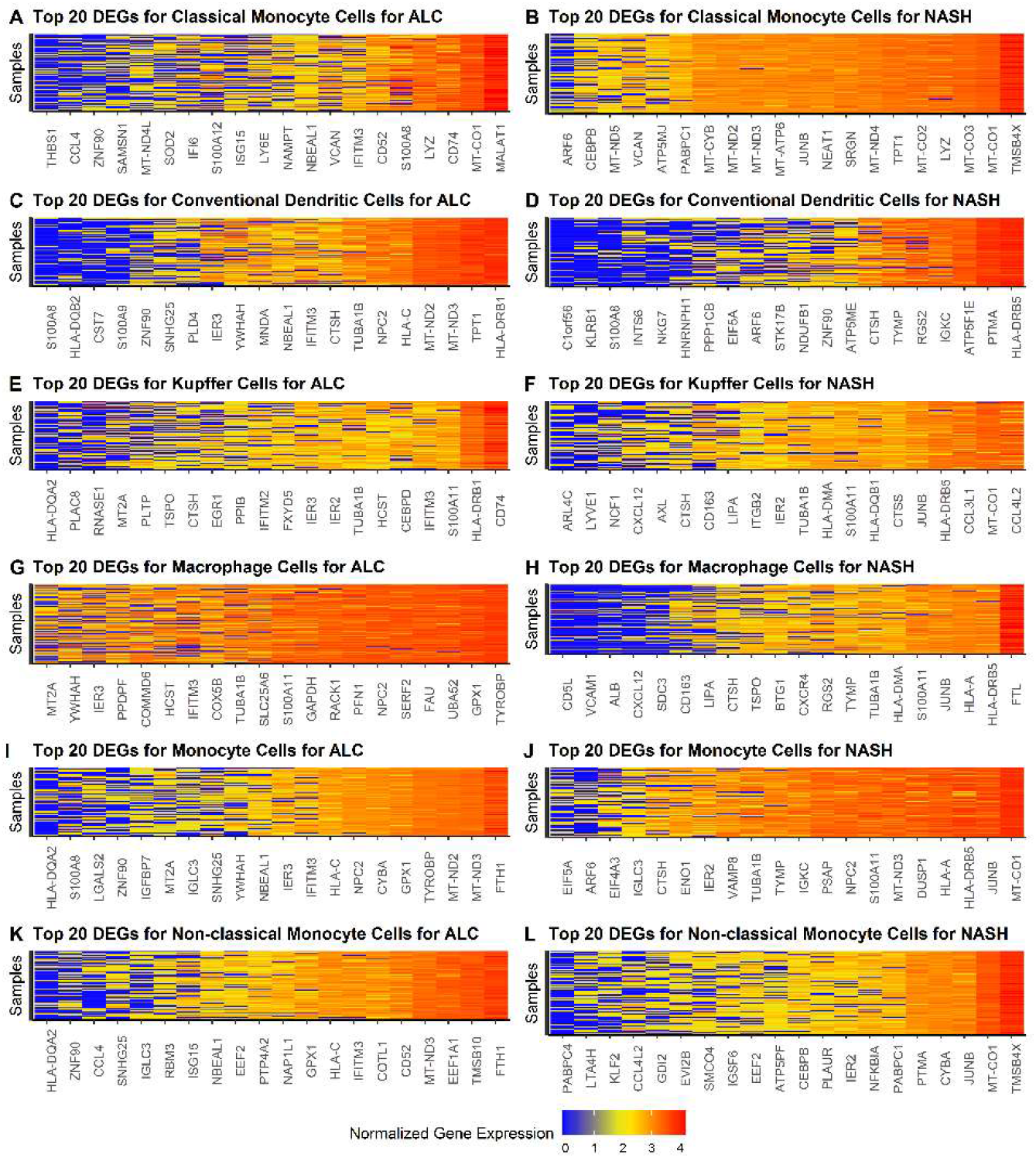
Heatmap of top 20 cell-type-specific DEGs: (**A**) DEGs for ALC tissues in classical monocyte cells. (**B)** DEGs for NASH tissues in classical monocyte cells. (**C**) DEGs for ALC tissues in conventional dendritic cells. (**D**) DEGs for NASH tissues in conventional dendritic cells. (**E**) DEGs for ALC tissues in Kupffer cells. (**F**) DEGs for NASH tissues in Kupffer cells. (**G**) DEGs for ALC tissues in macrophage cells. (**H**) DEGs for NASH tissues in macrophage cells. (**I**) DEGs for ALC tissues in monocyte cells. (**J**) DEGs for NASH tissues in monocyte cells. (**K**) DEGs for ALC tissues in non-classical monocyte cells. (**L**) DEGs for NASH tissues in non-classical monocyte cells. All DEGs are sorted and normalized. Upregulated genes are marked in red and downregulated genes are marked in blue. DEGs=differentially expressed genes. ALC=alcoholic liver cirrhosis. NASH=non-alcoholic steatohepatitis.

### Exploring Candidate Biomarker Genes: Master Regulator Analysis

Next, we used Master Regulator Analysis (MRA) to identify the top 5 master regulators of ALC and NASH using the DEG lists found in the previous step as the centroids. The individual master regulator genes for ALC (Figure 4A) and NASH (Figure 4B) are displayed in Figure 4. They depict the top five protein networks with the highest integrated normalized enrichment score and their 12 highest-likelihood network presumed targets. The MRA results reveal a strong shared up-regulation of HLA proteins, specifically HLA-DPA1, HLA-DRB1, HLA-DPB1, HLA-DQB1, and HLA-DRB5. In addition, our results show a unique up-regulation of interferon-induced proteins (i.e IFI44L and ISG15) and an invariant polypeptide of MHC class II antigen (CD74) for ALC and up-regulation of two regulators of actin polymerization (i.e., PFN1 and TMSB4X) for NASH. The full list of genes found in each master regulator network is presented in Table 1.

**Table 1.**
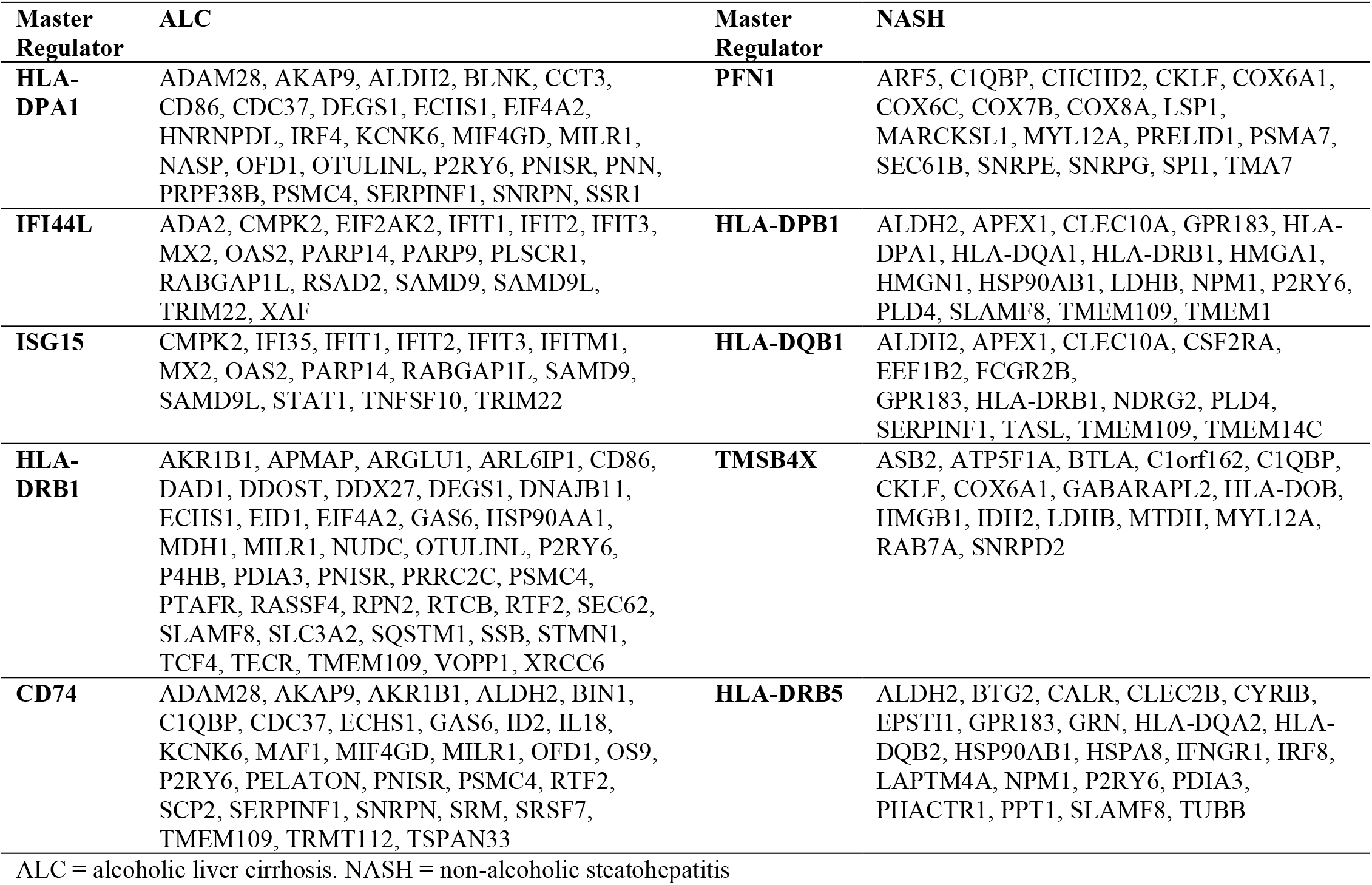
List of genes in each network for the top 5 master regulators.

**Figure 4.**
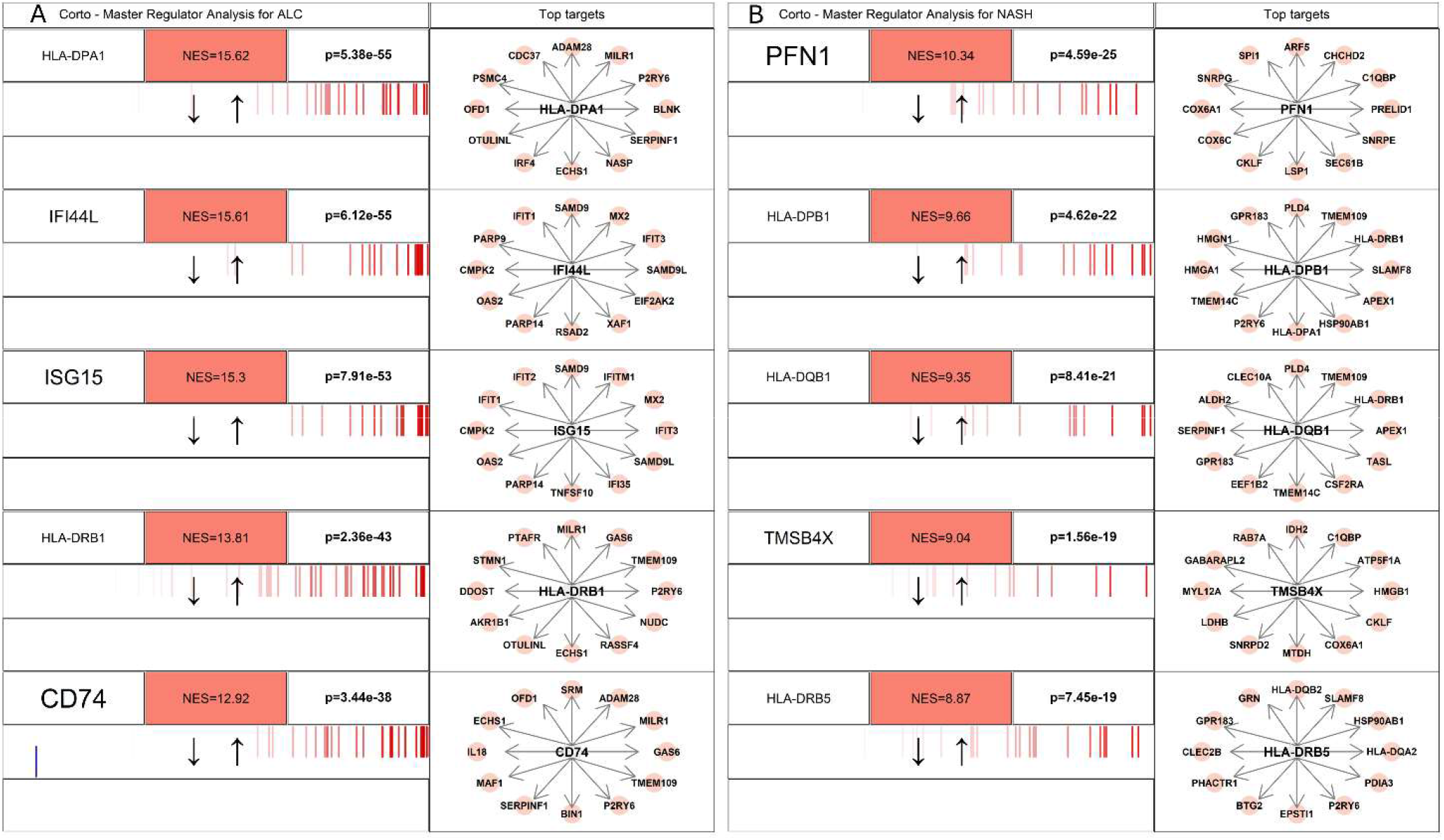
Master Regulator Genes for ALC and NASH based on the DEG Lists. The visualization was obtained through the *mraplot* function of the *R CRAN* package *corto*. For each analyzed network, the centroid is identified by DEGs for each condition. The genes in each network are displayed in a barcode-like diagram showing all genes based on their differential expression, from most downregulated (left) to most upregulated (right). Positively correlated targets are shown in red, while negatively correlated targets are shown in blue. Normalized Enrichment Score (NES) and p-value are also provided. Additionally, the 12 highest-likelihood network putative targets of each protein are displayed to the right. Upregulated targets are shown in red, downregulated targets in blue, with a pointed arrow indicating predicted activation by the centroid protein, and a blunt arrow indicating predicted repression. ALC=alcoholic liver cirrhosis. NASH=non-alcoholic steatohepatitis.

### Exploring Candidate Biomarker Genes: Weighted Key Driver Analysis (wKDA)

In addition to the MRA algorithm, we used wKDA to identify the key drivers of ALC and NASH conditions using the embedded Bayesian Liver Network in the Mergeomics online pipeline, resulting in the network neighbourhoods that have an over-representation of disease-associated genes. In ALC tissues, the top five key drivers include C1QA, HLA-DMB, RGS1, FCGR2A, and CD300A. Figure 5A shows the upregulation of C1QA and HLA-DMB in most of the samples and the down-regulation of RGS1, FCGR2A, and CD300A in ALC tissues. In NASH tissues, the top five key drivers are TYROBP, PTPRC, RAC2, NCKAP1L, and INPP5D. Figure 5B illustrates the upregulation of TYROBP and PTPRC in most of the samples and the downregulation of RAC2, NCKAP1L, and INPP5D in NASH tissues.

**Figure 5.**
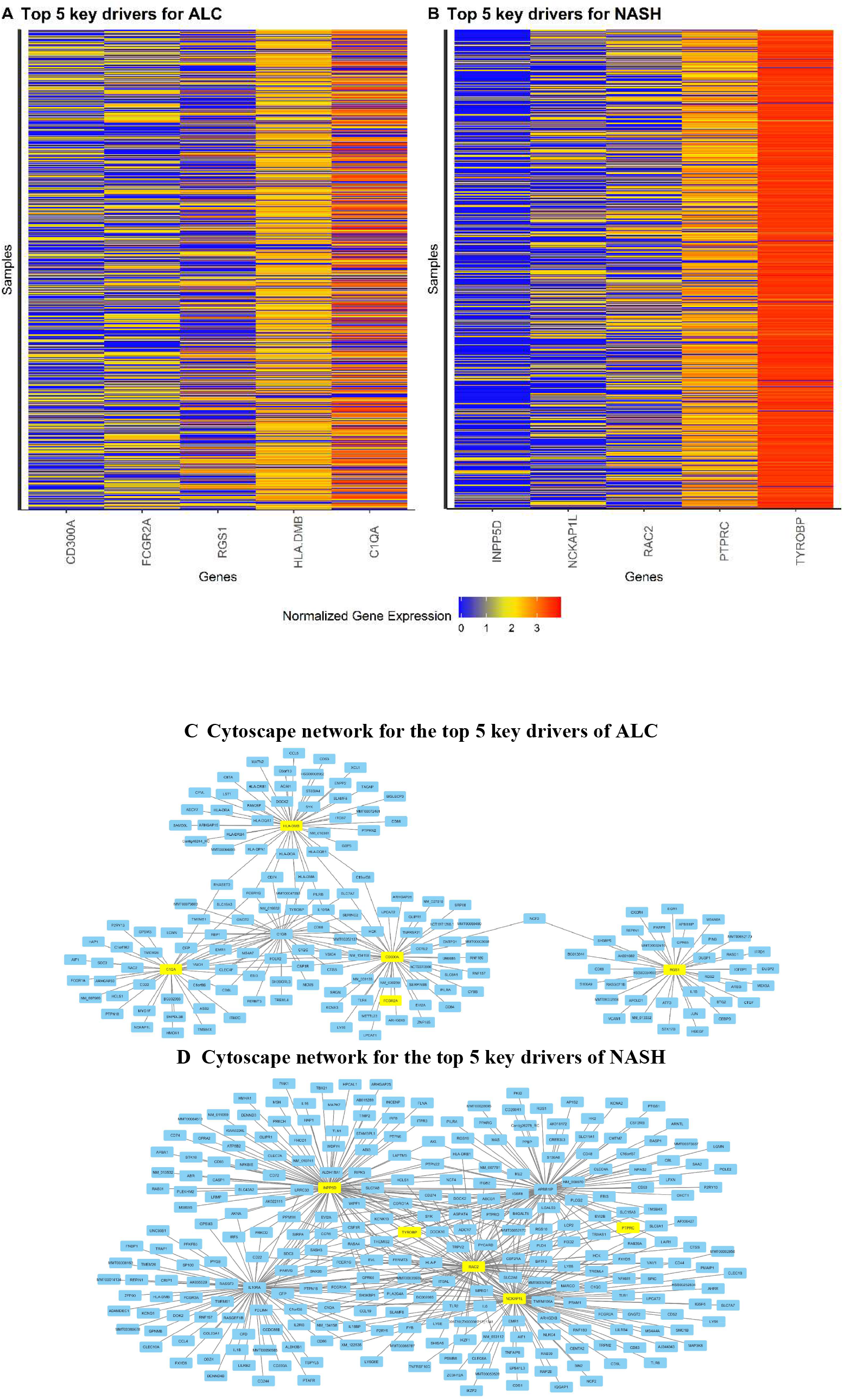
Heatmap of top five key drivers: (**A**) shows the top 5 key drivers for ALC tissues. (**B**) shows the top 5 key drivers for NASH tissues. **(C)** shows the network on all genes connected to the top 5 key drivers in ALC tissues. **(D)** shows the network on all genes connected to the top 5 key drivers in NASH tissues All genes are sorted and normalized. Upregulated genes are marked in red, and downregulated genes are marked in blue. ALC=alcoholic liver cirrhosis. NASH=non-alcoholic steatohepatitis.

## Discussion

Recent studies have increasingly utilized bioinformatic tools to investigate the mechanisms underlying liver diseases. In this study, we conducted DEG analysis along with two machine learning-based network algorithms, namely MRA and wkDA, and have identified key biomarkers associated with ALC and NASH using two publicly available RNA-seq datasets. Our findings reveal a notable upregulation of HLA proteins in both ALC and NASH. Furthermore, we identified IFI44L, ISG15, CD74, CD300A, C1QA, RGS1, and FCGR2A as potential key master regulators and biomarkers specifically for ALC. In contrast, PFN1, TMSB4X, TYROBP, PTPRC, RAC2, NCKAP1L, and INPP5D emerged as exclusive master regulators and biomarkers for NASH. We discussed the possible mechanism of these biomarkers in the following paragraphs.

### Shared Master Regulators in NASH and ALC

The MRA analysis highlights a significant upregulation of HLA proteins in both ALC and NASH tissues. These HLA proteins, part of the major histocompatibility complex (MHC) class II antigens, play a key role in immune regulation by presenting extracellular antigens to T-lymphocytes (27), triggering CD4+ T cell proliferation and B cell activation (28). Zhang Y et al. found a significant increase in MHC-II expression in human cirrhotic liver samples compared to non-cirrhotic samples (29). In particular, their study identified overexpression of MHC-II in endothelial cells with partial expression in liver sinusoidal endothelial cells. In our cell-type-specific DEG analysis, we observed upregulation of HLA-DRB1 in conventional dendritic cells and Kupffer cells associated with ALC, and upregulation of HLA-DRB5 in conventional dendritic cells, Kupffer cells, macrophages, and monocytes associated with NASH. In addition, our analysis showed that HLA-DRB1 and HLA-DRB5 upregulate the P2RY6 receptor, a factor implicated in the progression of both conditions (30,31).

Additionally, TMEM109, or mitsugumin 23, is differentially regulated in ALC and NASH by various HLA proteins. In NASH, its upregulation is controlled by HLA-DPB1 and HLA-DQB1, while in ALC, HLA-DRB1 regulates its expression. TMEM109, functioning as a voltage-gated channel, is involved in apoptosis and cellular responses to DNA damage, offering protection against oxidative stress, a common factor in liver diseases like ALD and NASH (32). Moreover, the protein αB-crystallin (hspb5) is upregulated as part of the liver’s defence mechanism, protecting cells from oxidative stress and apoptosis while stabilizing the cytoskeleton (33).

### Master Regulators and Key Drivers Exclusive to ALC

Our findings revealed the upregulation of interferon-induced proteins IFI44L and ISG15, along with the expression of the invariant polypeptide CD74, exclusively in ALC tissues. These proteins are central to type-I interferon signalling, which is activated in ALC and contributes to inflammatory responses. In support, Støy et al. and Stärkel et al. demonstrated reduced ISG15 expression in the liver and activation of the interferon signalling pathway in hepatocytes exposed to alcohol (34,35). Additionally, STAT3 activation as a downstream effect of type-I interferon signalling is implicated in liver fibrosis, a key feature of ALC progression (36,37). CD74, a receptor for macrophage migration inhibitory factor (MIF), plays a critical role in inflammatory pathways within the liver, with studies suggesting its inhibition could have therapeutic potential for ALC (38,39).

Our weighted Key Driver Analysis (wKDA) results identified HLA-DMB as a critical driver in ALC. We explored the role of HLA proteins in the immune response during inflammation in the previous section of the discussion. Another key driver of ALC, the complement C1qA chain (C1QA) encodes the A-chain polypeptide of serum complement subcomponent C1q, which interacts with C1r and C1s to form the initial component of the serum complement system. Shen et al. found elevated C1q levels in patients with autoimmune hepatitis, indicating ongoing inflammation (40). Similarly, Ren et al. showed upregulation of C1QA in monocytes and macrophages in ALC tissue compared to healthy controls (41). Additionally, RGS1, which modulates the GPCR signalling pathway, was identified as a key driver for ALC. Its downregulation in liver cirrhosis and its role in CD8 T-cell exhaustion have been documented in several studies (42–44).

Other crucial ALC drivers include FCGR2A (CD32) and CD300A. FCGR2A, an immunoglobulin receptor on immune cells, is important for cell recognition, phagocytosis, and cytotoxicity, as well as platelet function and the migration of inflammatory cytokines (45,46). The last top key driver of ALC, Human CD300A expressed on NK cells and subsets of T and B cells, modulates both innate and adaptive immunity by inhibiting Toll-like receptor (TLR) signalling pathways, which are vital for immune activation and inflammatory responses. CD300A also regulates leukocyte proliferation, differentiation, and immunity (47–54).

### Master Regulators and Key Drivers Exclusive to NASH

Our study demonstrated the up-regulation of two actin polymerization regulators, PFN1 and TMSB4X, in NASH tissues through the actin cytoskeleton pathway, which is commonly elevated in NAFLD (55). Cytoskeleton proteins, responsive to mitochondrial, ER, and oxidative stress, are implicated in NAFLD pathophysiology (55,56). Previous research has reported elevated cytoskeleton proteins in NASH biopsies (57–60), with PFN1 being notably up-regulated in hepatic stellate cells (HSCs), monocytes, and macrophages in liver fibrosis (57,58). TMSB4X, a variant of Thymosin beta 4, plays a significant role in liver fibrosis by influencing HSC activation and extracellular matrix production (61–63). Additionally, studies suggest that targeting cytoskeleton proteins could offer therapeutic benefits in NASH, with interventions such as Chinese herbal medicine and CD36 knockdown showing reductions in fibrosis markers (64,65)

The wKDA analysis of NASH tissues revealed five key drivers, including TYROBP, which is crucial for NK cell function by mediating receptor interactions and activating signalling pathways (66). Studies have shown TYROBP’s significant role in NAFLD, acting as a central hub gene in interaction networks, and its elevated presence in NASH models highlights its contribution to disease progression (67,68). Similarly, PTPRC (CD45), a vital regulator of immune responses, is up-regulated in NAFLD and linked to increased leukocyte infiltration in NASH (69). Research has shown PTPRC to be a hub gene in gene expression networks and a critical marker for immune activity during NASH progression (8).

Other top drivers include RAC2, which regulates IL-1β expression, with its deletion leading to increased RAC1 activity, contributing to NASH pathology by promoting cirrhosis and apoptosis (70– 72). Another top key driver of NASH, NCKAP1L (HEM1), predominantly expressed in immune cells, plays a key role in cell motility and immune cell migration, serving as a biomarker for NAFLD (73). Castro et al. provided a detailed report on NCKAP1L deficiency in humans, revealing its association with immunodeficiency, lymphoproliferation, and hyperinflammation, characterized by T cell exhaustion (74). Finally, INPP5D (SHIP1), a phosphatase regulating the PI3K/AKT pathway, is implicated in liver fibrosis and immune cell abundance (75–78). Its down-regulation is associated with a decrease in T cells and macrophages but an increase in NK cells, marking its role in NASH progression and immune regulation (79,80).

### Strength and Limitations

This study presented a comprehensive framework for analyzing single-cell RNA-seq data, demonstrating the potential of combining advanced network-based machine learning techniques with conventional differential gene expression (DEG) analysis to uncover actionable prognostic markers. Upon validation in clinical trials, these markers could profoundly enhance patient care by enabling more precise risk stratification and guiding the design of individualized treatment strategies. This approach offers the potential for early identification of high-risk patients, the selection of targeted therapies, and improvements in clinical outcomes while reducing treatment-related morbidity and optimizing healthcare resource utilization. The translation of these findings into routine clinical practice would mark a critical advancement in the pursuit of personalized medicine, particularly in the context of liver cancer.

Several limitations must be acknowledged in our study. First, our findings are based on gene expression and immune-related network analyses utilizing single-cell gene expression data. While scRNA-seq provides unprecedented resolution into the heterogeneity of individual cells, it has inherent limitations. These include potential biases introduced by dropout events, where genes may not be detected due to low expression levels, as well as technical variability that can affect the accuracy of cell-type identification and quantification. Additionally, the immune-related network analyses rely on inferred interactions, which may not fully capture the dynamic and context-specific nature of immune cell communication. Furthermore, our analysis does not account for potential spatial relationships between cells or the broader tissue microenvironment, which could influence immune cell behaviour and gene expression patterns. These factors may limit the generalizability of our results, particularly in more complex, multicellular systems. Second, while the results provide valuable insights, the specific contributions of these disease-related biomarkers to the pathogenesis remain unclear. Thus, additional experiments are required to elucidate the biological functions of these genes.

## Conclusion

This study introduced a comprehensive framework for single-cell RNA-seq data analysis, combining advanced network-based machine learning with traditional DEG analysis to identify actionable prognostic markers for ALC and NASH. Key findings revealed a pronounced upregulation of HLA proteins in both ALC and NASH tissues. Additionally, specific proteins were identified as exclusive master regulators and biomarkers: IFI44L, ISG15, CD74, CD300A, C1QA, RGS1, and FCGR2A for ALC, and PFN1, TMSB4X, TYROBP, PTPRC, RAC2, NCKAP1L, and INPP5D for NASH. These proteins hold promise as diagnostic markers and targets in drug development.

## Data Availability

The datasets analyzed in the current study are publicly available in the Gene Expression Omnibus (GEO) repository under accession numbers GSE136103 and GSE115469. These can be accessed via the following link: http://www.ncbi.nlm.nih.gov/geo

## Conflicts of interest

The work described herein represents the views of the authors and not of Applied Pharmaceutical Innovation or any of its funders.

## Notes

### Competing Interest Statement

The work described herein represent the views of the authors and not of the Applied Pharmaceutical Innovation or any of its funders.

### Funding Statement

This study did not receive any funding

