## Supplementary material for "Identification of Shared and Unique Key Biomarkers of Alcohol Liver Cirrhosis and Non-Alcoholic Steatohepatitis Through Machine Learning Network-Based Algorithms": Supplementary Material.pdf

### Supplementary Materials:

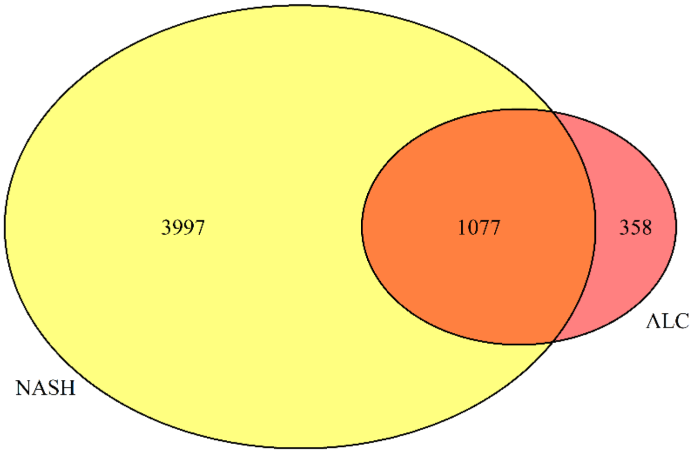


**Figure S1:** The number of unique and shared DEGs for ALC and NASH
ALC = alcoholic liver cirrhosis. NASH = non-alcoholic steatohepatitis

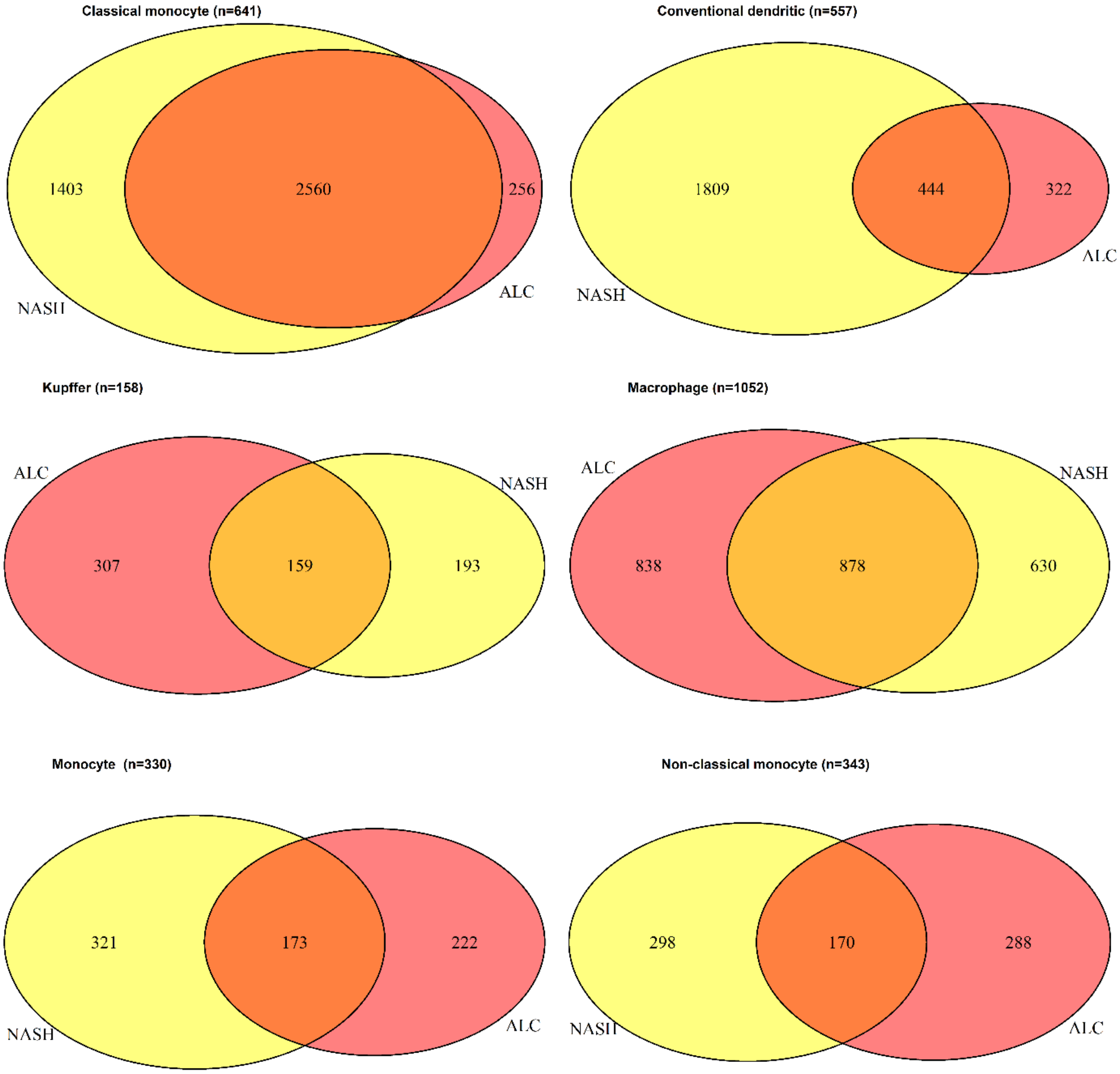


**Figure S2:** The number of unique and shared cell-type-specific DEGs for ALC and NASH
ALC = alcoholic liver cirrhosis. NASH = non-alcoholic steatohepatitis

**Table S1:** Top 20 cell-type-specific DEGs for ALC versus healthy and NASH versus healthy

| Cell Types | Top 20 DEGs ALC | Top 20 DEGs NASH | Shared |
|---|---|---|---|
| **Classical monocyte** | CD52,IFITM3,LYZ,NAMPT,CCL4,SOD2,CD74,S100A8,NBEAL1,S100A12,ZNF90,MALAT1,ISG15,VCAN,LY6E,MT-CO1,MT-ND4L,IFI6,SAMSN1,THBS1 | MT-CO1,MT-ND3,MT-ND4,MT-ND2,TPT1,MT-CO2,LYZ,MT-CO3,JUNB,MT-ATP6,MT-ND5,SRGN,PABPC1,MT-CYB,ATP5MJ,TMSB4X,VCAN,NEAT1,CEBPB,ARF6 | LYZ, VCAN, MT-CO1 |
| **Conventional dendritic** | HLA-C,ZNF90,NBEAL1,TPT1,NPC2,IER3,MT-ND2,MT-ND3,CTSH,HLA-DQB2,HLA-DRB1,MNDA,IFITM3,CST7,YWHAH,S100A9,TUBA1B,PLD4,S100A8,SNHG25 | HLA-DRB5,IGKC,CTSH,C1orf56,HNRNPH1,ATP5ME,STK17B,EIF5A,INTS6,NDUFB1,ARF6,RGS2,S100A8,NKG7,PPP1CB,ATP5F1E,PTMA,ZNF90,TYMP,KLRB1 | ZNF90, CTSH, S100A8, |
| **Kupffer** | MT2A,CD74,IER3,HLA-DQA2,IFITM3,IER2,S100A11,IFITM2,PLAC8,TUBA1B,HLA-DRB1,PPIB,CTSH,PLTP,RNASE1,FXYD5,CEBPD,HCST,EGR1,TSPO | LIPA,CCL3L1,HLA-DRB5,JUNB,TUBA1B,CCL4L2,S100A11,IER2,CXCL12,HLA-DMA,CTSH,CD163,MT-CO1,NCF1,AXL,HLA-DQB1,CTSS,ITGB2,ARL4C,LYVE1 | S100A11, TUBA1B, CTSH |
| **Macrophage** | S100A11,IER3,GPX1,TUBA1B,IFITM3,TYROBP,RACK1,FAU,SLC25A6,PFN1,SERF2,PPDPF,HCST,NPC2,YWHAH,COMMD6,MT2A,GAPDH,UBA52,COX5B | JUNB,S100A11,LIPA,FTL,RGS2,HLA-DRB5,CD163,CD5L,CTSH,VCAM1,TUBA1B,HLA-A,BTG1,TYMP,CXCR4,SDC3,CXCL12,ALB,TSPO,HLA-DMA | S100A11, TUBA1B |
| **Monocyte** | MT-ND2,MT-ND3,IFITM3,ZNF90,HLA-C,LGALS2,NPC2,HLA-DQA2,TYROBP,IER3,NBEAL1,MT2A,FTH1,SNHG25,IGLC3,IGFBP7,S100A8,GPX1,YWHAH,CYBA | MT-CO1,IGKC,HLA-DRB5,JUNB,DUSP1,CTSH,NPC2,S100A11,EIF5A,PSAP,ENO1,MT-ND3,EIF4A3,VAMP8,HLA-A,IGLC3,TYMP,IER2,TUBA1B,ARF6 | MT-ND3, IGLC3 |
| **Monocyte non-classical** | TMSB10,PTP4A2,EEF1A1,EEF2,CCL4,FTH1,IFITM3,GPX1,NBEAL1,CD52,NAP1L1,HLA-DQA2,MT-ND3,COTL1,ZNF90,HLA-C,IGLC3,ISG15,SNHG25,RBM3 | EEF2,NFKBIA,PABPC1,TMSB4X,PTMA,MT-CO1,JUNB,CEBPB,IGSF6,GDI2,CYBA,LTA4H,SMCO4,CCL4L2,KLF2,PABPC4,PLAUR,IER2,EVI2B,ATP5PF | -- |

DEG = differentially expressed genes. ALC = alcoholic liver cirrhosis. NASH = non-alcoholic steatohepatitis
